# A Fully Automated Deep Learning-based Network For Detecting COVID-19 from a New And Large Lung CT Scan Dataset

**DOI:** 10.1101/2020.06.08.20121541

**Authors:** Mohammad Rahimzadeh, Abolfazl Attar, Seyed Mohammad Sakhaei

## Abstract

COVID-19 is a severe global problem, and AI can play a significant role in preventing losses by monitoring and detecting infected persons in early-stage. This paper aims to propose a high-speed and accurate fully-automated method to detect COVID-19 from the patient’s CT scan images. We introduce a new dataset that contains 48260 CT scan images from 282 normal persons and 15589 images from 95 patients with COVID-19 infections. At the first stage, this system runs our proposed image processing algorithm to discard those CT images that inside the lung is not properly visible in them. This action helps to reduce the processing time and false detections. At the next stage, we introduce a novel method for increasing the classification accuracy of convolutional networks. We implemented our method using the ResNet50V2 network and a modified feature pyramid network alongside our designed architecture for classifying the selected CT images into COVID-19 or normal with higher accuracy than other models. After running these two phases, the system determines the condition of the patient using a selected threshold. We are the first to evaluate our system in two different ways. In the single image classification stage, our model achieved 98.49% accuracy on more than 7996 test images. At the patient identification phase, the system correctly identified almost 234 of 245 patients with high speed. We also investigate the classified images with the Grad-CAM algorithm to indicate the area of infections in images and evaluate our model classification correctness.

## 1 Introduction

On January 30, 2020, the World Health Organization(WHO) announced the outbreak of a new viral disease as an international concern for public health, and on February 11, 2020, WHO named of the disease caused by the new coronavirus: COVID-19 [31]. The first patients with COVID-19 were observed in Wuhan, China. These people were associated with the local wild animal market, which indicates the possibility of transmitting the virus from animals to humans [28]. The severe outbreak of the new coronavirus spread rapidly throughout China and then spread to other countries. The virus disrupted many political, economic, and sporting events and affected the lives of many people worldwide.

The most important feature of the new coronavirus is it’s fast and wide-spreading capability. The virus is mainly transmitted directly from people with the disease to others; It is transmitted indirectly through the surfaces and air in the environment in which the infected people come in contact with it [31]. As a result, correctly identifying the symptoms of people with the disease and quarantining them plays a significant role in preventing the disease.

New coronavirus causes viral pneumonia in the lungs, which results in severe acute respiratory syndrome. The new coronavirus causes a variety of changes in the sufferer. The most common symptoms of new coronavirus are fever, dry cough, and tiredness [31]. The symptoms of this disease vary from person to person [19]. Other symptoms such as loss of sense of smell and taste, headache, and sore throat may occur in some patients, but severe symptoms that indicate the further progression of COVID-19 include shortness of breath, chest pain, and loss of ability to move or Talking [31].

There are several methods for definitive diagnosis of COVID-19, including reverse transcriptase-polymerase chain reaction (RT-PCR), Isothermal nucleic amplification test, Antibody test, Serology tests, and medical imaging [32].

RT-PCR is the primary method of diagnosing COVID-19 and many viral diseases. However, the method is restricted for some of the assays as higher expertise and experimentation are required to develop new assays [8]. Besides, the lack of diagnostic kits in most contaminated areas around the world is leading researchers to come up with new and easier ways to diagnose the disease.

Due to the availability of medical imaging devices in most treatment centers, the researchers analyze CT scans and X-rays to detect COVID-19. In most patients with COVID-19, infections are found in the lungs of people with new coronavirus that can help diagnose the disease. Analysis of CT scans of patients with COVID-19 showed pneumonia caused by the new coronavirus [28]. With the approval of radiologists for the ability to use CT scans and X-rays to detect COVID-19, various methods have been proposed to use these images.

Most patients who have COVID-19 symptoms at least four days later have X-rays and CT scans of their lungs, showing infections that confirm the presence of a new coronavirus in their body [3]. Although medical imaging is not recommended for definitive diagnosis, it can be used for early COVID-19 diagnosis due to the limitations of other methods [2].

In [33, 3], some patients with early-onset COVID-19 symptoms were found to have new coronavirus infections on their CT scans. At the same time, their RT-PCR test results were negative, then both tests were repeated several days later, and RT-pCR confirmed the CT scan’s diagnostic results. Although medical imaging is not recommended for the definitive diagnosis of COVID-19, it can be used as a primary diagnostic method for the COVID-19 to quarantine the Suspicious person and prevent the virus from being transmitted to others in the early stages of the disease.

The advantage of using medical imaging is the ability to visualize viral infections by machine vision. Machine vision has many different methods, one of the best of which is deep learning [10]. Machine vision and deep learning have many applications in medicine [26], agriculture [22], economics [9], etc., which have eliminated human errors and created automation in various fields.

The use of machine vision and deep learning is one of the best ways to diagnose tumors and infections caused by various diseases. This method has been used for various medical images, such as segmentation of lesions in the brain and skin [18], Applications to Breast Lesions, and Pulmonary Nodules [4], sperm detection and tracking [24] and state-of-the-art bone suppression in x-rays images[34].

On the other hand, diagnosing the disease by computer vision and deep learning is much more accurate than radiologists. For example, in [12], the accuracy of the method used is about 90%, while the accuracy of radiologists’ diagnosis is approximately 70%. Due to the effectiveness of machine vision and deep learning in medical imaging, especially CT scan and X-ray images, machine vision and deep learning have been used to diagnose COVID-19.

In this paper, we introduce a fully-automated method for detecting COVID-19 cases from the output files(images) of the lung HRCT scan device. This system does not need any medical expert for system configuration and takes all the CT scans of a patient and clarifies if that patient is infected to COVID-19 or not.

We also introduce and share a new dataset that we called COVID-CTset that contains 15589 COVID-19 images from 95 patients and 48260 normal images from 282 persons. At the first stage of our work, we apply an image processing algorithm for selecting those CT Scan images that inside the lung, and the possible infections are observable in them. In this way, we speed up the process because the network does not have to analyze all the images. Also, we improve the accuracy by giving the network only the proper images.

After that, we will train and test three deep convolutional neural networks for classifying the selected images. One of them is our proposed enhanced convolutional model, which is designed to improve classification accuracy. At the final stage, we evaluate our fully automated system in two different ways. The first way is single image classification, which is evaluated on more 7996 images, and the second is the fully automated system that we evaluated on almost 245 patients and 41892 images.

We also investigate the infected areas of the COVID-19 classified images by segmenting the infections using a feature visualization algorithm.

The general view of our work in this paper is represented in fig. 1.

**Figure 1:**
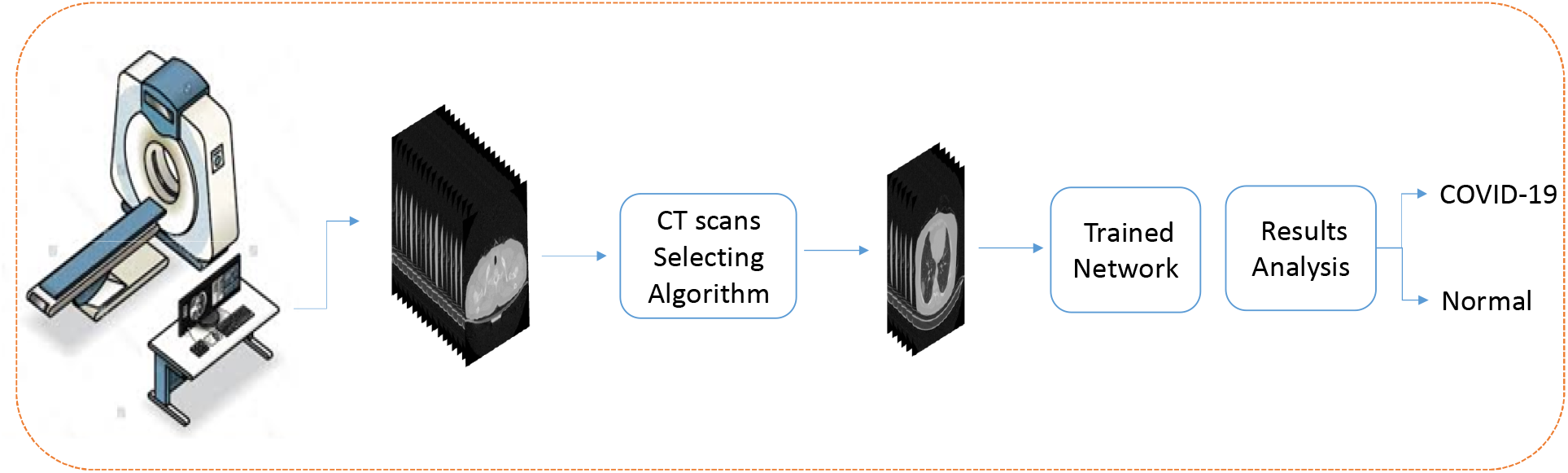
General view of our proposed method for automated patients classification.

In [21,17], using existing deep learning networks, they have identified COVID-19 on chest X-ray images and introduced the network with high accuracy. In [23], by concatenating Xception and Resnet50v2 networks and using chest X-ray images, they were able to diagnose normal patients, pneumonia, and COVID-19, with an overall accuracy of 99.5 and 91.4 in the COVID-19 class, which was evaluated on 11302 images.

In [14], 3322 eligible CT scans were selected from the 3506 CT scans of different persons and used to learn and evaluate the proposed network, COVNet. In another study, CT scans of 120 people (2482 CT scans) were collected, half of which (60 people) were COVID-19, and classified by different networks, which was the most accuracy equal to 97.38% [27].

In [12], CT scans of 287 patients were collected, including three classes of COVID-19, Community-acquired pneumonia (CAp), or other viral diseases in the lungs, and other diseases or healthy, and then, using the innovative algorithm called CovidCTNet, to classify the data with 90% accuracy.

In [30], CT scans of 5372 patients have been collected in several hospitals in China, which have been used in learning and evaluating the presented Innovative Deep Learning Network to classify data into three classes. In [29], CT scans have been used to segment infections caused by the new coronavirus.

The rest of the paper is organized as follows: In section 2, we will describe the dataset, neural networks, and proposed algorithms. In section 3, the experimental results are presented, and in section 4, the paper is discussed. In section 5, we have concluded our paper, and in the end, the links to the shared codes and dataset are provided.

## 2 Materials and methods

### 2.1 COVID-CTset

COVID-CTset^1^ is our introduced dataset. It was gathered from Negin radiology located at Sari in Iran between March 5th to April 23rd, 2020. This medical center uses a SOMATOM Scope model and syngo CT VC30-easyIQ software version for capturing and visualizing the lung HRCT radiology images from the patients. The format of the exported radiology images was 16-bit grayscale DICOM format with 512*512 pixels resolution. As the patient’s information was accessible via the DICOM files, we converted them to TIFF format, which holds the same 16-bit grayscale data but does not conclude the patients’ private information. Also, this format is easier to use with standard programming libraries. In the addressed link^2^ at the end of this paper, the general information (age, sex, time of radiology imaging) for each patient is available.

One of our novelties is using a 16bit data format instead of converting it to 8bit data, which helps improve the method’s results. Converting the DICOM files to 8bit data may cause losing some data, especially when few infections exist in the image that is hard to detect even for clinical experts. This lost data may be the difference between different images or the values of the pixels of the same image. The pixels’ values of the images differ from 0 to almost 5000, and the maximum pixels values of the images are considerably different. So scaling them through a consistent value or scaling each image based on the maximum pixel value of itself can cause the mentioned problems and reduce the network accuracy. So each image of COVID-CTset is a TIFF format, 16bit grayscale image.

In some stages of our work, we used the help of clinical experts under the supervision of the third author, a radiology specialist, to separate those images that the COVID-19 infections are clear. To make these images visible with standard monitors, we converted them to float by dividing each image’s pixel value by the maximum pixel value of that image. This way, the output images had a 32bit float type pixel values that could be visualized by standard monitors, and the quality of the images was good enough for analysis. Some of the images of our dataset are presented in fig. 2.

**Figure 2:**
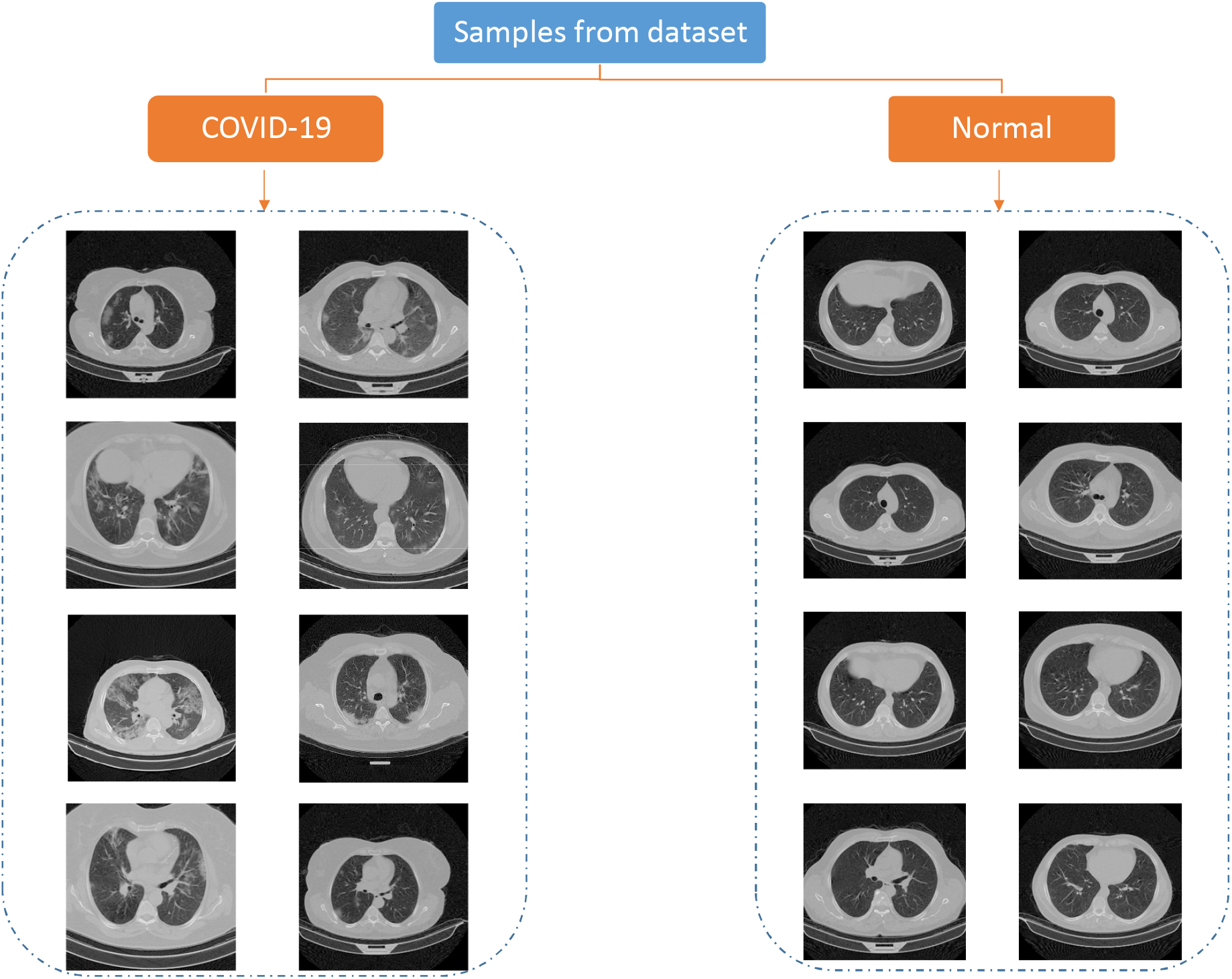
Some of the images in COVID-CTset

COVID-CTset is made of 15589 images that belong to 95 patients infected to COVID-19 and 48260 images of 282 normal people (table 1). Each patient has three folders that each folder includes the CT scans captured from the CT imaging device with a different thickness.^3^

**Table 1:** COVID-CTset data distribution

|  | COVID-19 Patients | Normal Patients | COVID-19 Images | Normal Images |
| --- | --- | --- | --- | --- |
| Number | 95 | 282 | 15589 | 48260 |

The distribution of the patients in COVID-CTset is shown in fig. 3

**Figure 3:**
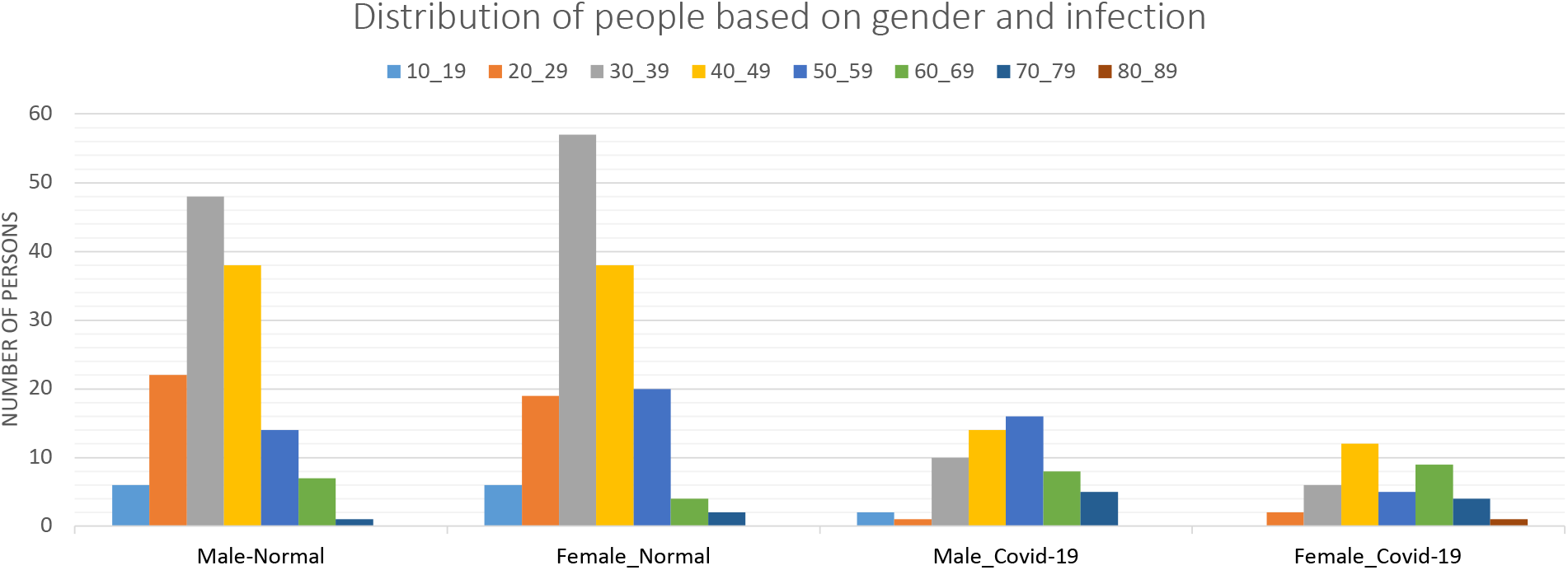
This figure shows the number of patients based on age, gender and infections.

### 2.2 CT Scans Selection Algorithm

The lung HRCT scan device takes a sequence of consecutive images(we can call it a video or consecutive frames) from the chest of the patient that wants to check his infection to COVID-19. In an image sequence, the infection points may appear in some images and not be shown in other images.

The clinical expert analyzes theses consecutive images and, if he finds the infections on some of them, indicates the patient as infected.

Many previous methods selected an image of each patient’s lung HRCT images and then used them for training and validation. Here we decided to make the patient lung analysis fully automated. Consider we have a neural network that is trained for classifying CVOID-19 cases based on a selected data that inside the lung was obviously visible in them. If we test that network on each image of an image sequence the belongs to a patient, the network may fail. Because at the beginning and the end of each CT scan image sequence, the lung is closed as it is depicted in fig. 4. Hence, the network has not seen these cases while training; it may result in wrong detections, and so does not work well.

**Figure 4:**
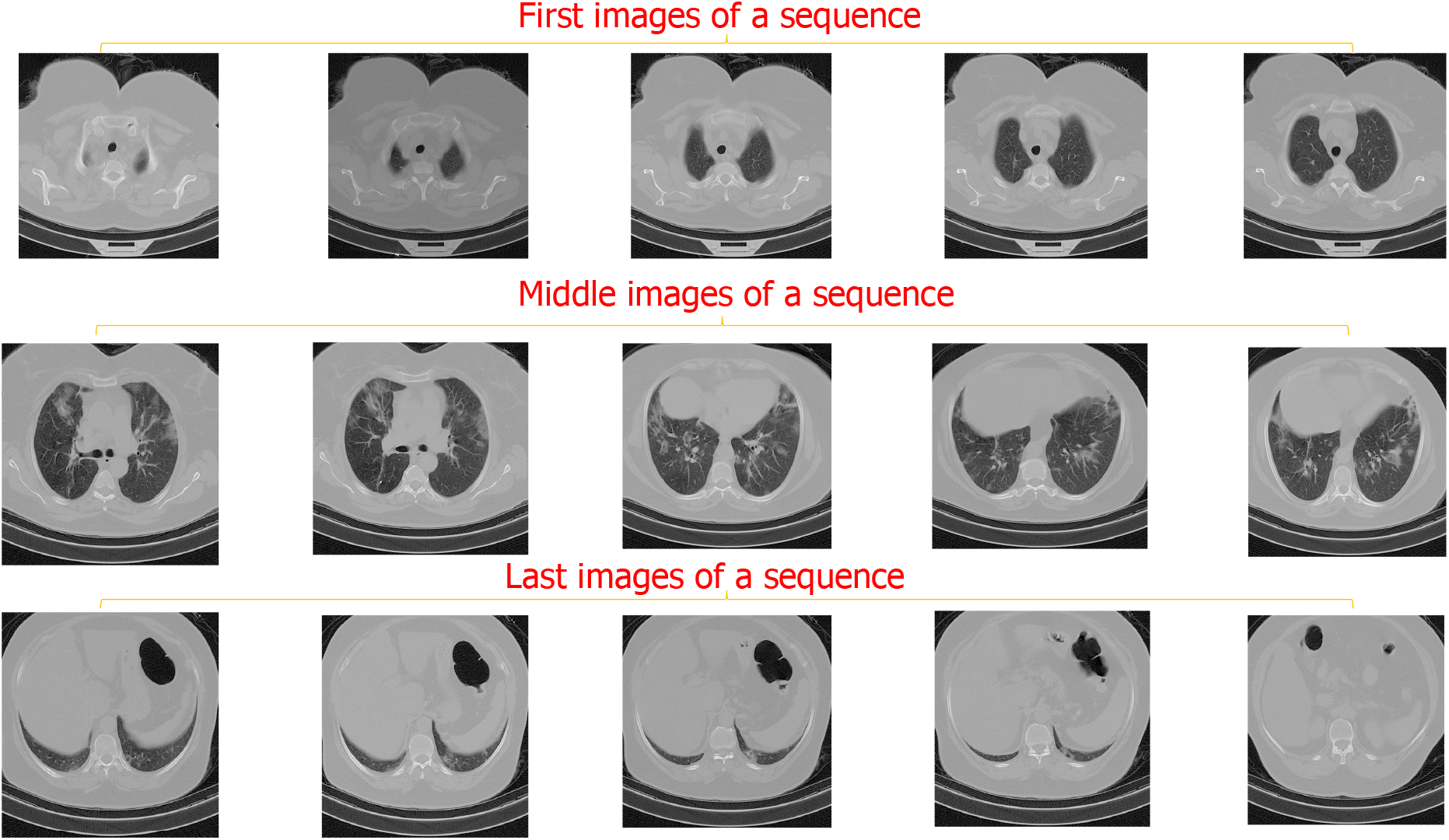
This figure presents some of the first, middle, and final images of a patient CT scans sequence. It is obvious from the images that in the first and the last images, inside the lung is not observable.

**Figure 5:**
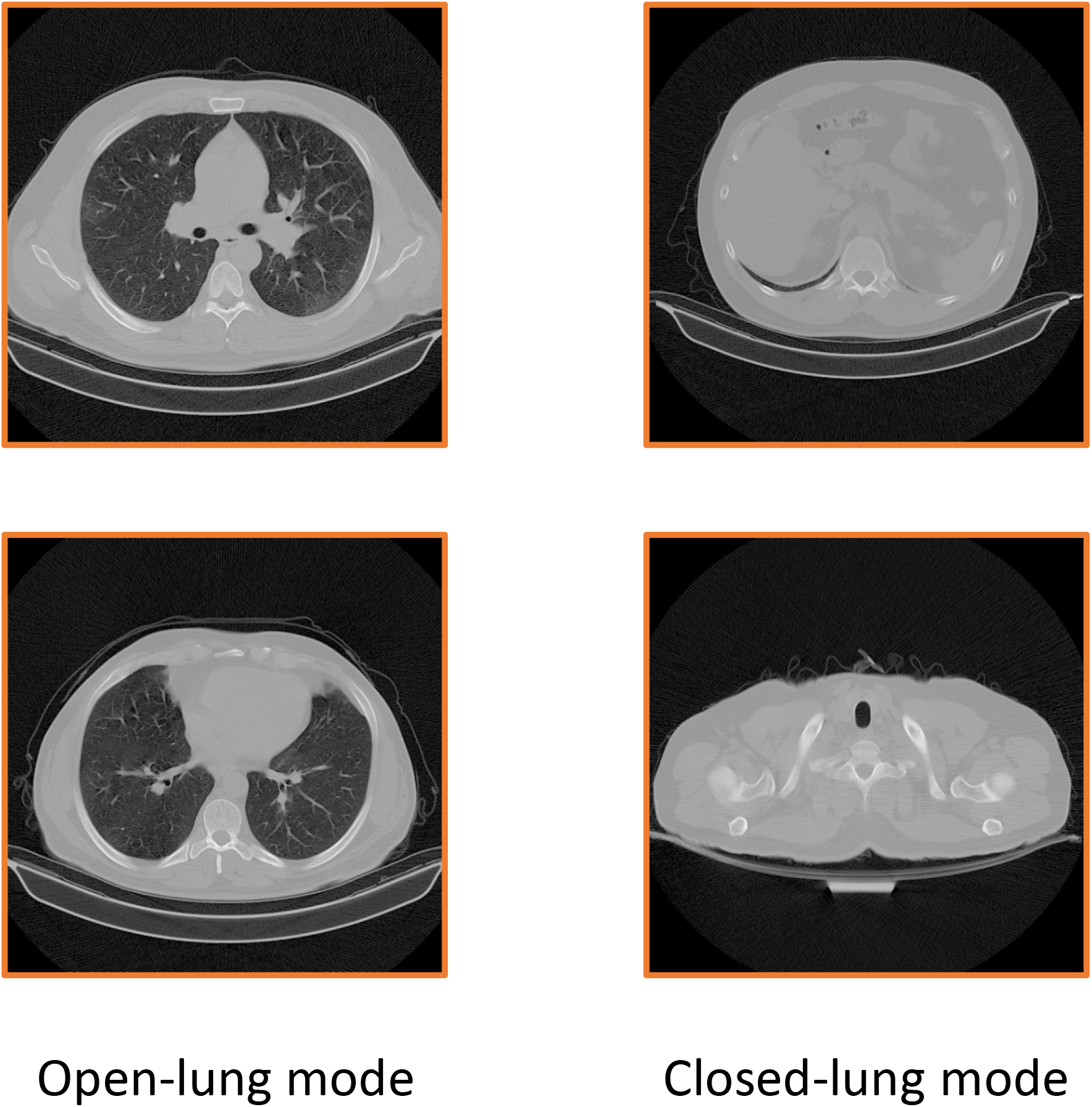
It can be visualized from this figure that a closed-lung has higher pixels values in the middle of the image.

To solve this, we can separate the dataset into three classes: infection-visible,no-infection, and lung-closed. Although this removes the problem but dividing the dataset into three classes has other costs like spending some time for making new labels, changing the network evaluating way. Also, it increases the processing time because the network shall see all the images of patient CT scans. But we propose some other techniques to discard the images that inside the lungs are not visible in them. Doing this also reduces performing time for good because, in the last method, the networks should have seen all the images, and now it only sees some selected images.

Fig. 6 shows the steps of the image-selection algorithm. As it is evident from fig. 5, the main difference between an open lung and closed lung is that the open lung image has lower pixel values(near to black) in the middle of the lung. First, we set a region in the middle of the images for analyzing the pixel values in them. This region should be at the center of the lung in all the images, so open-lung and closed-lung show the differences in this area. Unfortunately, the images of the dataset were not on one scale, and the lung’s position differed for different patients; so after experiments and analysis, as the images have 512*512 pixels resolution, we set the region in the area of 120 to 370 pixels in the x-axis and 240 to 340 pixels in the y-axis ([120,240] to [370,340]). This area shall justify in containing the information of the middle of the lung in all the images. Fig. 7 shows the selected region in some different images.

**Figure 6:**
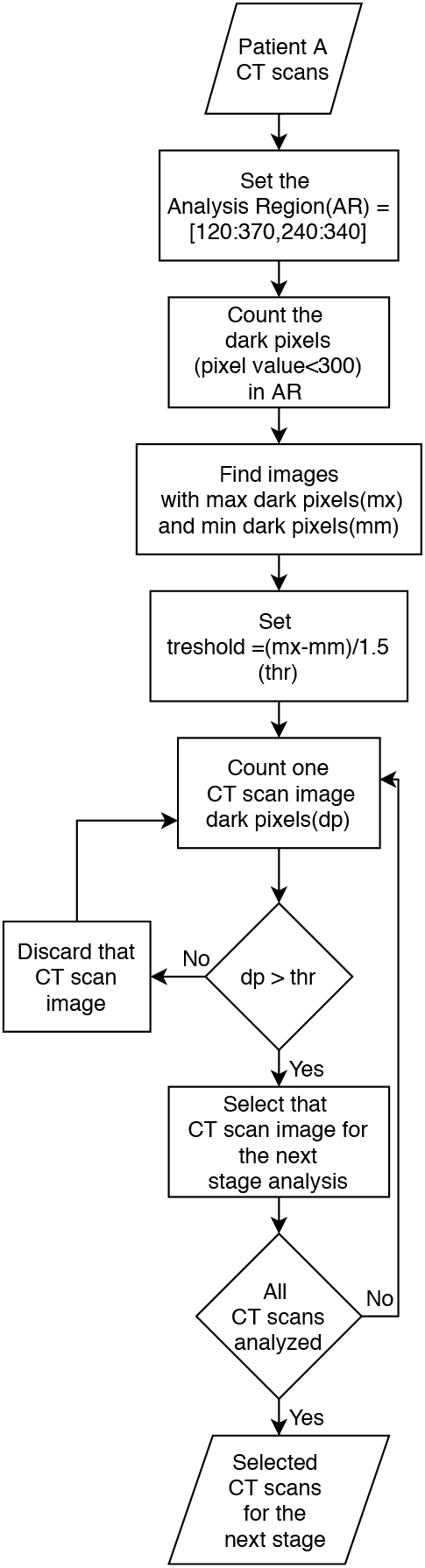
The flowchart of the proposed algorithm for selecting the efficient CT scan images of a sequence

**Figure 7:**
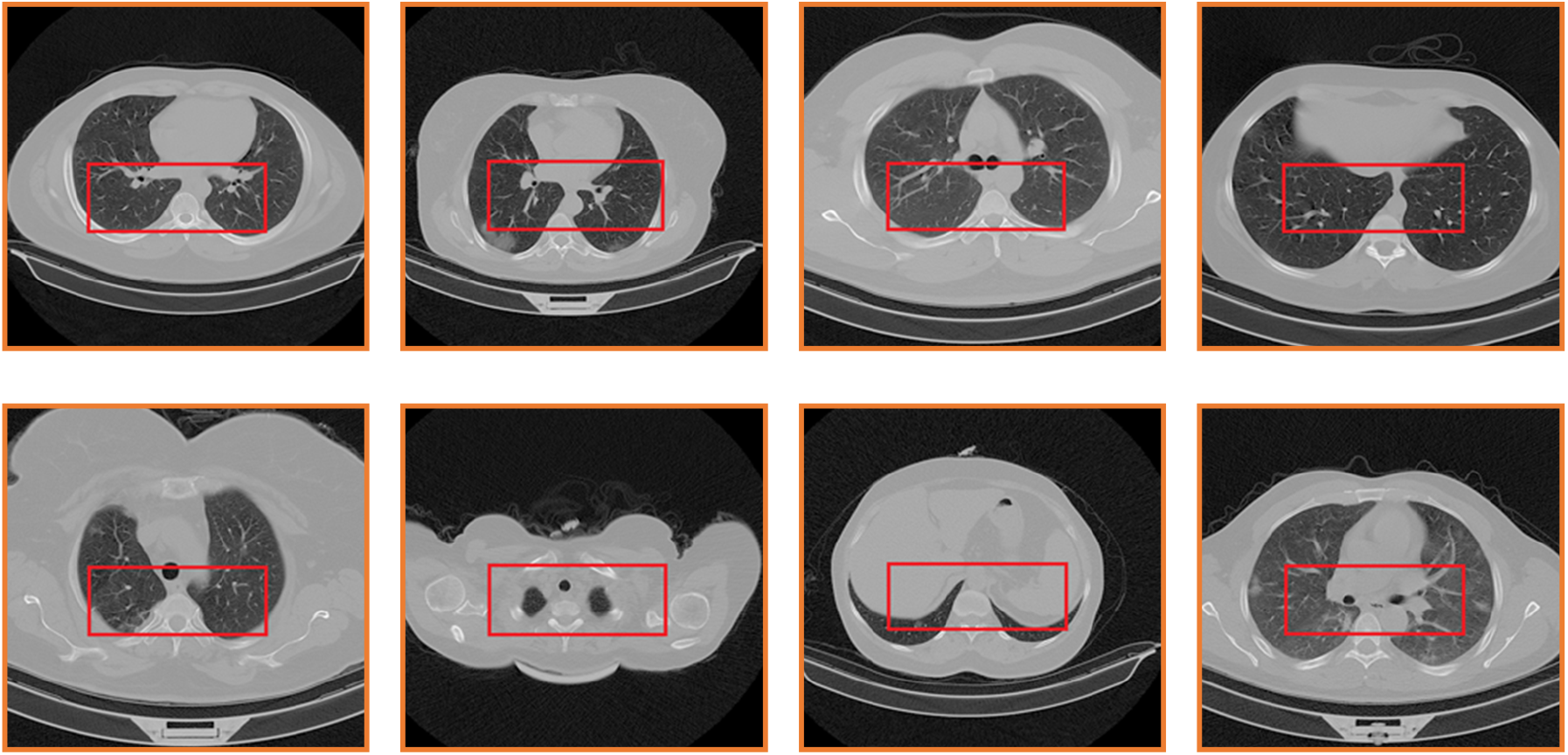
The selected region in different images with different scales

The images of our dataset are 16-bit grayscale images. The maximum pixel value between all the images is almost equal to 5000. This maximum value differs very much between different images. At the next step for discarding some images and selecting the rest of them from an image sequence that belongs to a patient, we aim to measure the pixels of each image in the indicated region that have less value than 300, which we call dark pixels. This number was chosen out of our experiments.

For all the images in the sequence, we count the number of pixels in the region with less value than 300. After that, we would divide the difference between the maximum counted number and the minimum counted number by 1.5. This calculated number is our threshold. For example, if a CT scan image sequence of a patient has 3030 pixels with a value of less than 300 in the region, and another has 30 pixels less than 300, the threshold becomes 2000. The image with less dark pixels in the region than the threshold is the image that the lung is almost closed in that, and the image with more dark pixels is the one that inside the lung is visible in it.

We calculated this threshold in this manner that the images in a sequence (CT scans of a patient) be analyzed together because, in one sequence, the imaging scale does not differ. After that, we discard those images that have less counted dark pixels than the calculated threshold. So the images with more dark pixels than the computed threshold will be selected to be given to the network for classification.

In fig. 8, the image sequence of one patient is depicted, where it can be observed, which of the images the algorithm discards and which will be selected.

**Figure 8:**
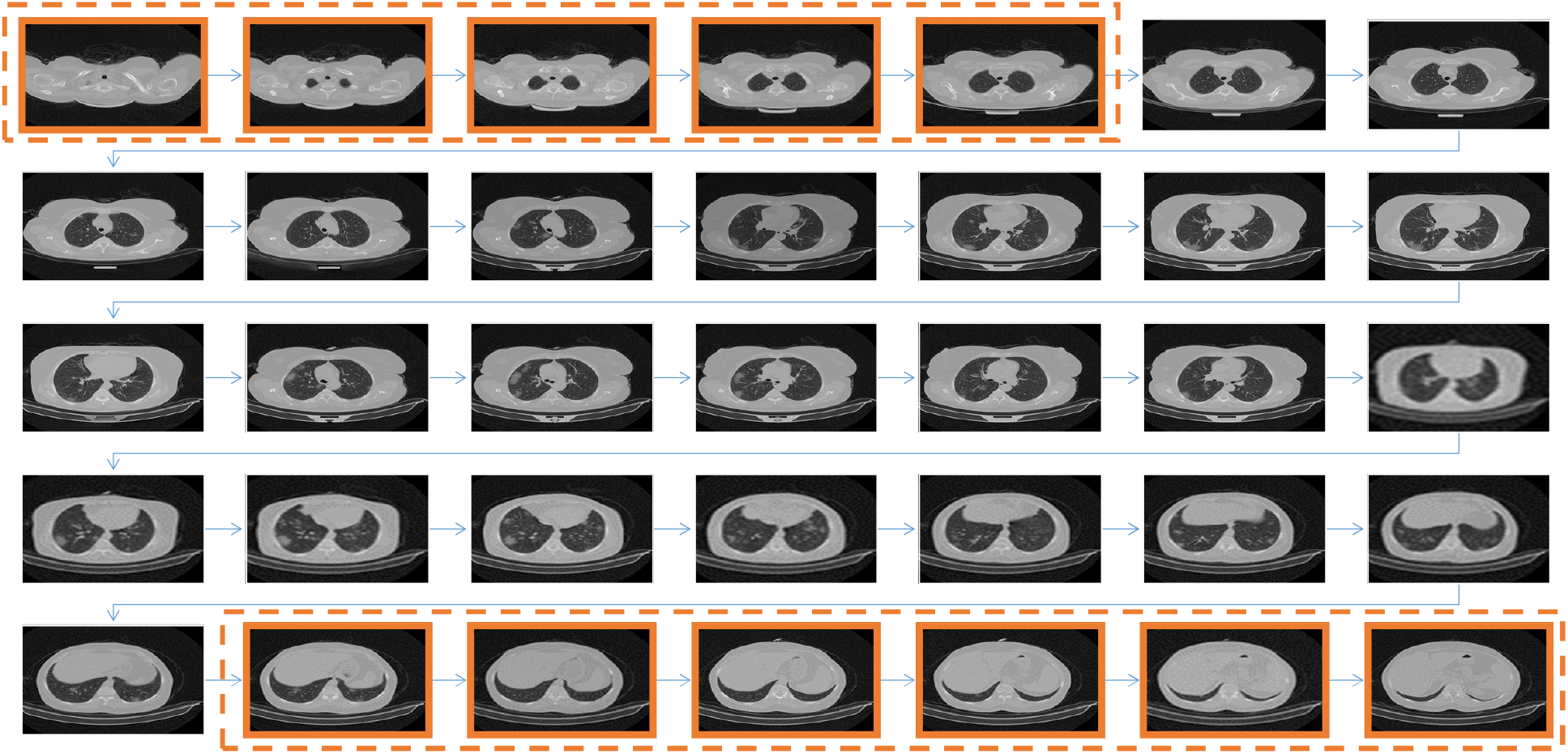
The CT scan images of a patient are shown in this figure. The highlighted images are the ones that the algorithm discards. It is observable that those images that clearly show inside the lung are selected to be classified at the next stage.

### 2.3 Enhanced deep convolutional neural network for classification

Machine Vision has been a superior method for advancing many fields like Agriculture [22], biomedical engineering[24, 20], industry [13] and others. Implementing machine vision methods on the deep neural networks, especially using the convolution layers, has resulted in extremely accurate performing. In this research, we used deep convolution networks to classify the selected CT scan images exported from the CT scan selecting algorithm into normal or COVID-19. We trained, evaluated, and compared three different deep convolutional networks: Xception [5], ResNet50V2 [11], and our proposed model.

Xception introduced new inception modules constructed of depth-wise, separable convolution layers (depth-wise convolutional layers followed by a point-wise convolution layer). Xception achieved one of the best results on ImageNet [7] dataset. ResNet50V2, is a upgraded version of ResNet50 [10]. In this neural network, the authors made some changes in the connections and skip-connections between blocks and increased network performance in the ImageNet dataset.

Feature pyramid network(FPN) was introduced by paper [15] and was utilized in RetinaNet [16] for enhancing object detection. FPN helps the network better learning and detecting objects at different scales that exist in an image. Some of the previous methods worked by giving an image pyramid (that includes different scales of the input image) to the network as the input. Doing this indeed improves the feature extraction process but also increases the processing time and is not efficient.

FpN solves this problem by generating a bottom-up and a top-down feature hierarchy with lateral connections from the network’s generated features at different scales. This helps the network generate more semantic features, so using FPN helps increase detection accuracy when there are objects with various scales in the image while not changing detection speed.

Although FPN was developed for object detection networks, we propose a new model that utilizes FPN for image classification. COVID-19 infections exist in different scales, so using FPN helps to extract various semantic features of the input image. Our model can detect COVID infections even when they are tiny and, more importantly, detect COVID false positives fewer because it learns better about the infection points.

In fig. 9, the architecture of the proposed network can be obsereved. We used ResNet50V2 as the backbone network and compared our model with ResNet50V2 and Xception. Other researchers can also use other models as the backbone. The FPN we used is like the original version of FPN [15] with this difference that we used concatenation layers instead of adding layers inside the feature pyramid network due to the authors’ experience.

**Figure 9:**
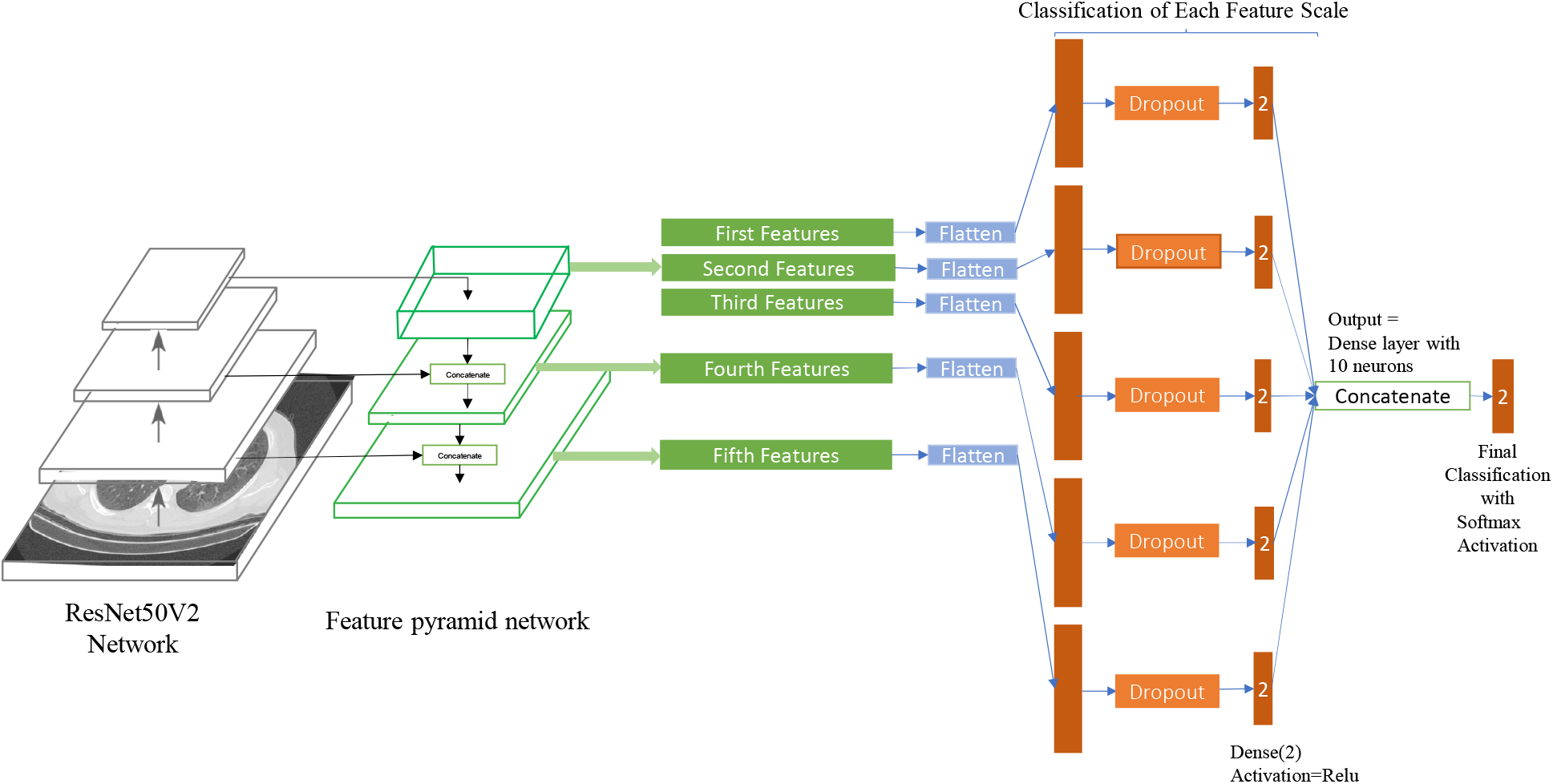
This figure shows our model, which uses ResNet50V2 as the backbone and applies the feature pyramid network and the designed layers for classification.

FPN extracts five final features that each one presents the input image features on a different scale. After that, we implemented dropout layers (to avoid overfitting), followed by the first classification layers. Note that we did not use softmax functions for the first classification layers because we wanted to feed them to the final classification layer, and as the softmax function computes each output neuron based on the ratio of other output neurons, it is not suitable for this place. Relu activation function is more proper.

At the end of the architecture, we concatenated the five classified layers (each consisting of two neurons) and made a ten neurons dense layer. Then we connected this layer to the final classification layer, which applies the softmax function. With running this procedure, the network utilizes different classification results based on various scales features. As a result, the network would become able to classify the images better.

Researchers can use our proposed model for running classification in other cases and datasets to improve classification results.

### 2.4 Training Phase

Our dataset is constructed of two sections. The first section is the raw data for each person that is described in section 2.1. The second section includes training, validation, and testing data. We converted the images to 32-bit float types on the TIFF format so that we could visualize them with standard monitors. Then we took the help of the clinical experts under the supervision of the third author(Radiology Specialist) in the Negin radiology center to select the infected patients’ images that the infections were clear on them. We used these data for training and testing the trained networks.

To report more real and accurate results, we separated the dataset into five folds for training, validation, and testing. Almost 20 percent of the patients with COVID19 were allocated for testing in each fold. The rest were considered for training, and part of the testing data has been considered for validating the network after each epoch while training. Because the number of normal patients and images was more than the infected ones, we almost chose the number of normal images equal to the COVID-19 images to make the dataset balanced. Therefore the number of normal images that were considered for network testing was higher than the training images. The details of the training and testing data are reported in table 2.

**Table 2:** Training and testing details of COVID-CTset

|  | Training Set |  |  |  | Testing Set |  |  |  |
| --- | --- | --- | --- | --- | --- | --- | --- | --- |
|  | COVID-19 Patients | COVID-19 Images | Normal Patients | Normal Images | COVID-19 Patients | COVID-19 Images | Normal Patients | Normal Images |
| Fold1 | 77 | 1820 | 45 | 1916 | 18 | 462 | 237 | 7860 |
| Fold2 | 72 | 1817 | 37 | 1898 | 23 | 465 | 245 | 7878 |
| Fold3 | 77 | 1836 | 53 | 1893 | 18 | 446 | 229 | 7883 |
| Fold4 | 81 | 1823 | 76 | 1920 | 14 | 459 | 206 | 7856 |
| Fold5 | 73 | 1832 | 71 | 1921 | 22 | 450 | 211 | 7785 |

From the information in table 2, the question may arise as to why the number of normal persons in the training set is less than the number of COVID-19 patients. Because in each image sequence of a patient with COVID-19, we allocated some of them with observable infections for training and testing. So the number of images for a COVID-19 patient is less than the number of images for a normal person.

We selected enough number of normal patients somehow that the number of normal images becomes almost equal to the number of COVID-19 class images. This number was enough for the network to learn to classify the images correctly, and the achieved results were high. As we had more normal images left, We selected a large number of normal data for testing so that the actual performance of our trained networks be more clear.

We trained our dataset on Xception[5], Resnet50V2[11] and our model until 50 epochs. For training the networks, we used transfer learning from the ImageNet [7] pre-trained weights to make the networks’ convergence faster. We chose the Nadam optimizer and the Categorical Cross-entropy loss function. We also used data augmentation methods to make learning more efficient and stop the network from overfitting.

It is noteworthy that we did not resize the images for training or testing so as not to lose the small data of the infections. Our training parameters are listed in table 3.

**Table 3:** Training Parameters

| Training Parameters | Xception | ResNet50V2 | Our model |
| --- | --- | --- | --- |
| Learning Rate | 1e-4 | 1e-4 | 1e-4 |
| Batch Size | 14 | 14 | 14 |
| Optimizer | Nadam | Nadam | Nadam |
| Loss Function | Categorical Crossentropy | Categorical Crossentropy | Categorical Crossentropy |
| Epochs | 50 | 50 | 50 |
| Horizontal/Vertical flipping | Yes | Yes | Yes |
| Zoom Range | 5% | 5% | 5% |
| Rotation Range | 0 - 360 degree | 0 - 360 degree | 0 - 360 degree |
| Width / Height Shifting | 5% | 5% | 5% |
| Shift Range | 5% | 5% | 5% |

As is evident from table 3, we used the same parameters for all the networks.

## 3 Experimental results

In this section, we report the results into two sections. The Image classification results section includes the results of the trained networks on the test set images. The Patient identification section reports the results of the automated system for identifying each person as normal or COVID-19.

We implemented our algorithms and networks on Google Colaboratory Notebooks, which allocated a Tesla P100 GPU, 2.00GHz Intel Xeon CPU, and 12GB RAM on Linux to us. We used Keras library [6] on Tensorflow backend [1] for developing and running the deep networks.

### 3.1 Image classification results

We trained each network on the training set with the explained parameters in section 2.4. We also used the accuracy metric while training for monitoring the network validation result after each epoch to find the training network’s best-converged version.

We evaluated the trained networks using four different metrics for each of the classes and the overall accuracy for all the classes as follows:

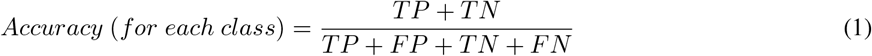

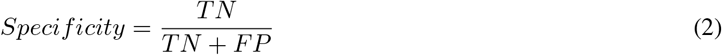

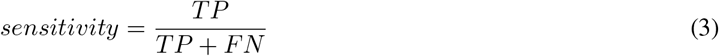

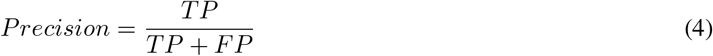

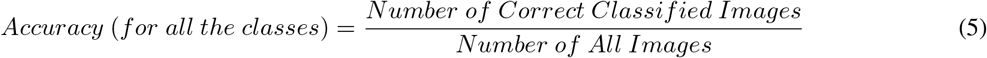

In these equations, for each class, *TP* (True Positive) is the number of correctly classified images, *FP* (False Positive) is the number of the wrong classified images, *FN* (False Negative) is the number of images that have been detected as a wrong class, and *TN* (True Negative) is the number of images that do not belong to another class and have not been classified as that class.

The results for each fold is reported in table 8. We also showed the average results between five folds in confusion matrices in fig. 10.

**Figure 10:**
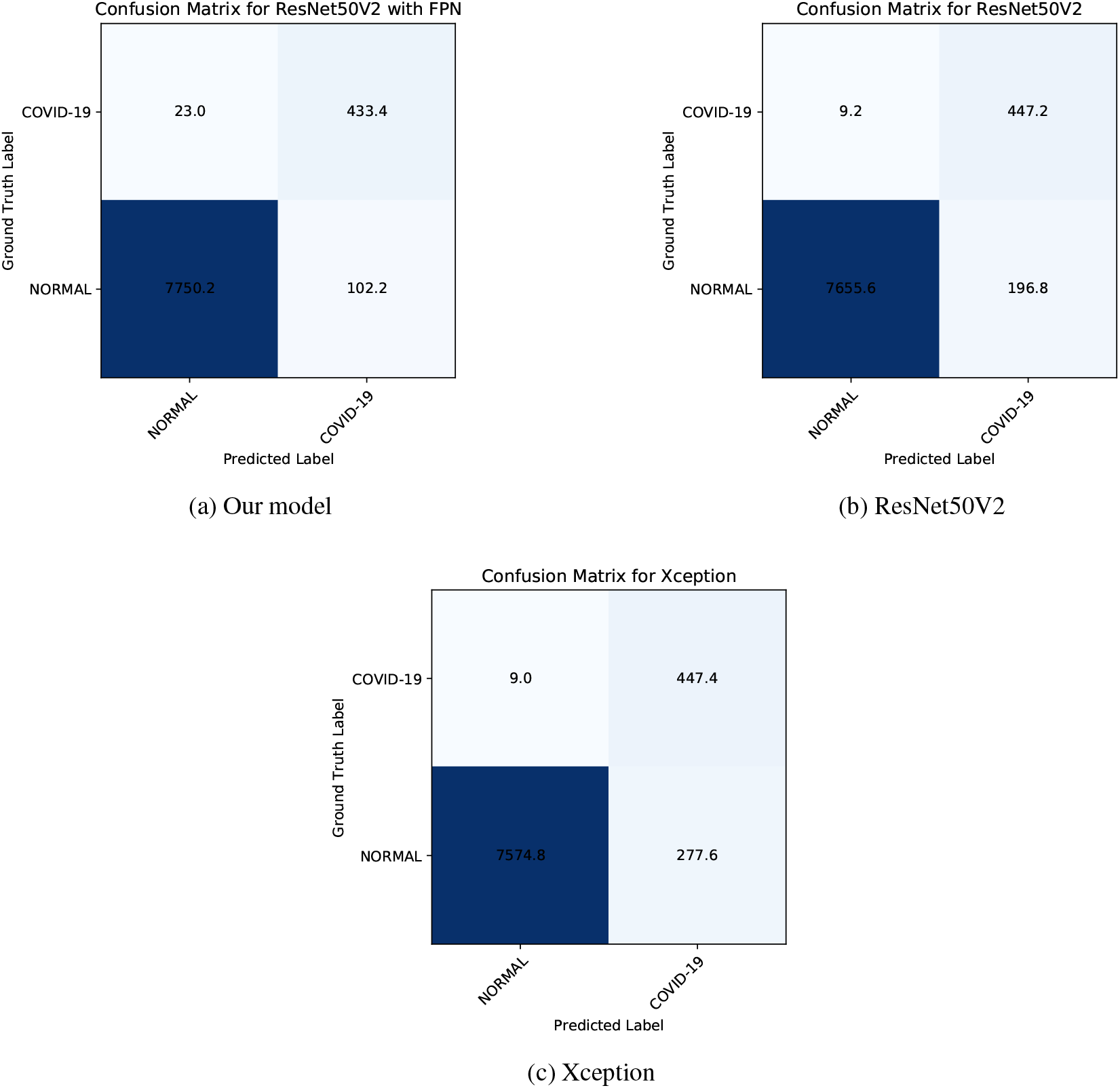
The average data between five folds are shown on these confusion matrices

Fig. 11 shows the training and validation accuracy in 20 epochs of the training procedure. Our model converges faster to higher accuracy. This is the ability of our model, which enhances the base model.

**Figure 11:**
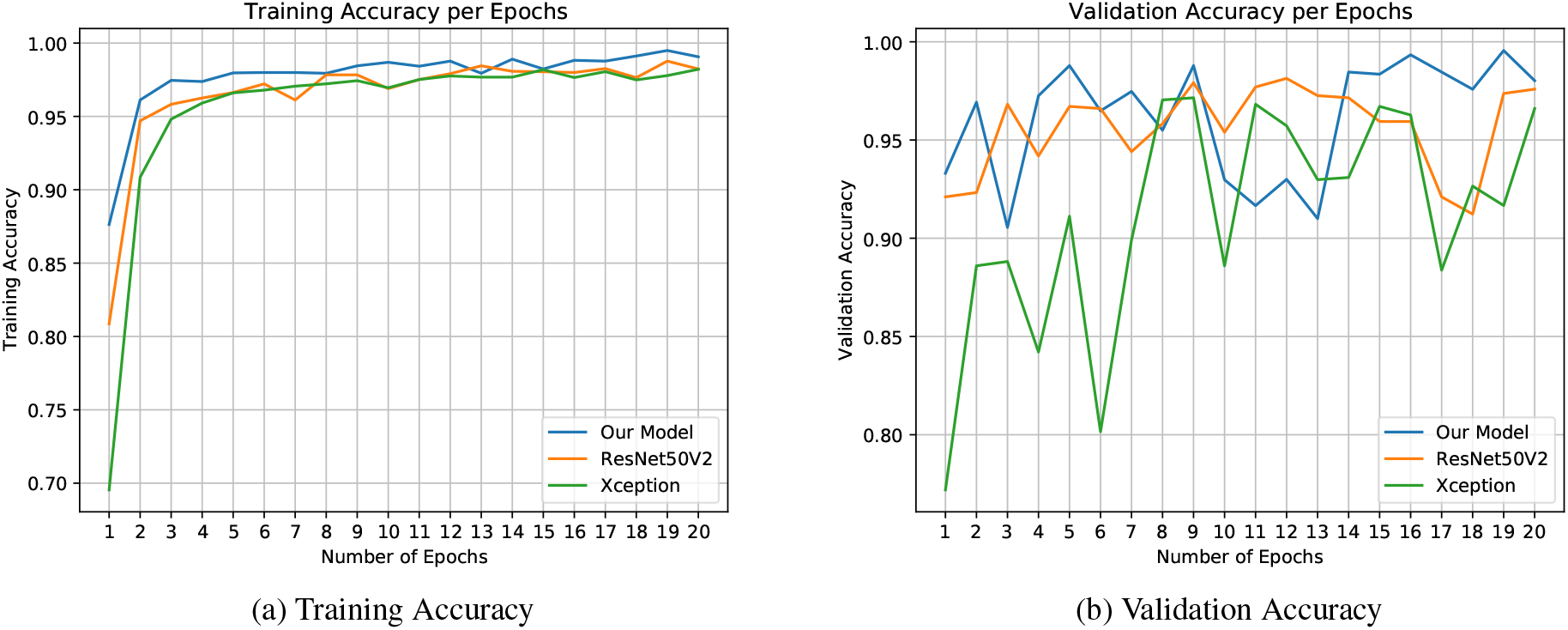
These plots show training and validation accuracy in 20 epochs. It is clear that our model is converging faster and reaches to higher values of accuracy.

### 3.2 Patient identification

In this section, we present the main results of our work. CT scan data is not like many other data like X-ray images, which can be evaluated by investigating one single image. CT scans are sequences of consecutive images (like videos), so for medical diagnosis, the system or the expert person must analyze more than one image.

Based on this condition, for proposing an automatic diagnosis system, the developers must evaluate their system differently than single image classification. As we know, until today, we are the first that have evaluated our model in this way.

If our proposed fully-automated system wants to check the infection of COVID-19 for a patient, it takes all the images of the patient CT scans as input. Then it processes them with the proposed CT scan selection algorithm to select the CT scans that the lung is visible in them. Those chosen images will be fed to the deep neural network to be classified as COVID-19 or normal.

For indicating the condition of a patient, we must set a threshold. For each patient, if the number of CT scan images, which are identified as COVID-19, be more than the threshold, that patient would be considered as infected; otherwise, his condition would be normal. This threshold value depends on the precision of the model. In trained models with high accuracy, the threshold can be set to zero, meaning if at least one CT scan image of a patient (between the filtered CT scans by the selection algorithm) is detected as COVID-19, that patient would be considered being infected.

The number of data used for evaluating our system in this section is listed in the table. 4.

**Table 4:** Details of the dataset used for the patient identification evaluation stage.

| Fold | Number of Images belonging to patients with COVID | Number of patients with COVID | Number of Images belonging to normal persons | Number of normal patients |
| --- | --- | --- | --- | --- |
| Fold1 | 2858 | 18 | 40094 | 237 |
| Fold2 | 3540 | 23 | 39302 | 245 |
| Fold3 | 3640 | 18 | 39391 | 229 |
| Fold4 | 2769 | 14 | 37323 | 206 |
| Fold5 | 2783 | 22 | 37758 | 211 |

Table 5 shows an interesting result. In this table, threshold 1 is zero, and threshold 2 is one-tenth of the filtered CT scan images. It means that after filtering the CT scan images by the selection algorithm, in threshold1, if at least one CT scan image be identified as COVID, the patient would be selected as infected. In threshold 2, if at least one-tenth of the number of the filtered CT scan images are detected as COVID, then the system determines that patient, infected to COVID-19.

**Table 5:** Comparison between different patient identification thresholds for each of the trained networks in fold1. Threshold 1 is zero and Threshold2 is equal to 0.1 of the CT images.

|  | Our model |  |  |  | ResNet50V2 |  |  |  | Xception |  |  |  |
| --- | --- | --- | --- | --- | --- | --- | --- | --- | --- | --- | --- | --- |
|  | COVID correct detected | Wrong detected as Normal | Normal correct detected | Wrong detected as COVID | COVID correct detected | Wrong detected as Normal | Normal correct detected | Wrong detected as COVID | COVID correct detected | Wrong detected as Normal | Normal correct detected | Wrong detected as COVID |
| Threshold1 | 17 | 1 | 231 | 6 | 17 | 1 | 205 | 32 | 17 | 1 | 195 | 42 |
| Threshold2 | 16 | 2 | 232 | 5 | 17 | 1 | 222 | 15 | 17 | 1 | 221 | 16 |

With these circumstances, in table 5, we can see that our model performs very well in threshold 1, and this shows the high performance of this network. But other networks do not execute well in threshold 1, which indicates that these networks’ accuracy is lower than our model. This remarkable result is caused by the feature pyramid network that gives a high ability to detect the infections correctly and not to detect false points as infections.

The users can select this threshold based on their model accuracy. In the following (6), we used the second threshold (equal to one-tenth) for reporting the full results. But we recommend using the first threshold for accurate models like our model.

The results of Patient identification for each of the trained networks in each fold are available in table 6. The speed of the fully automated system is reported in table 7.

**Table 6:**
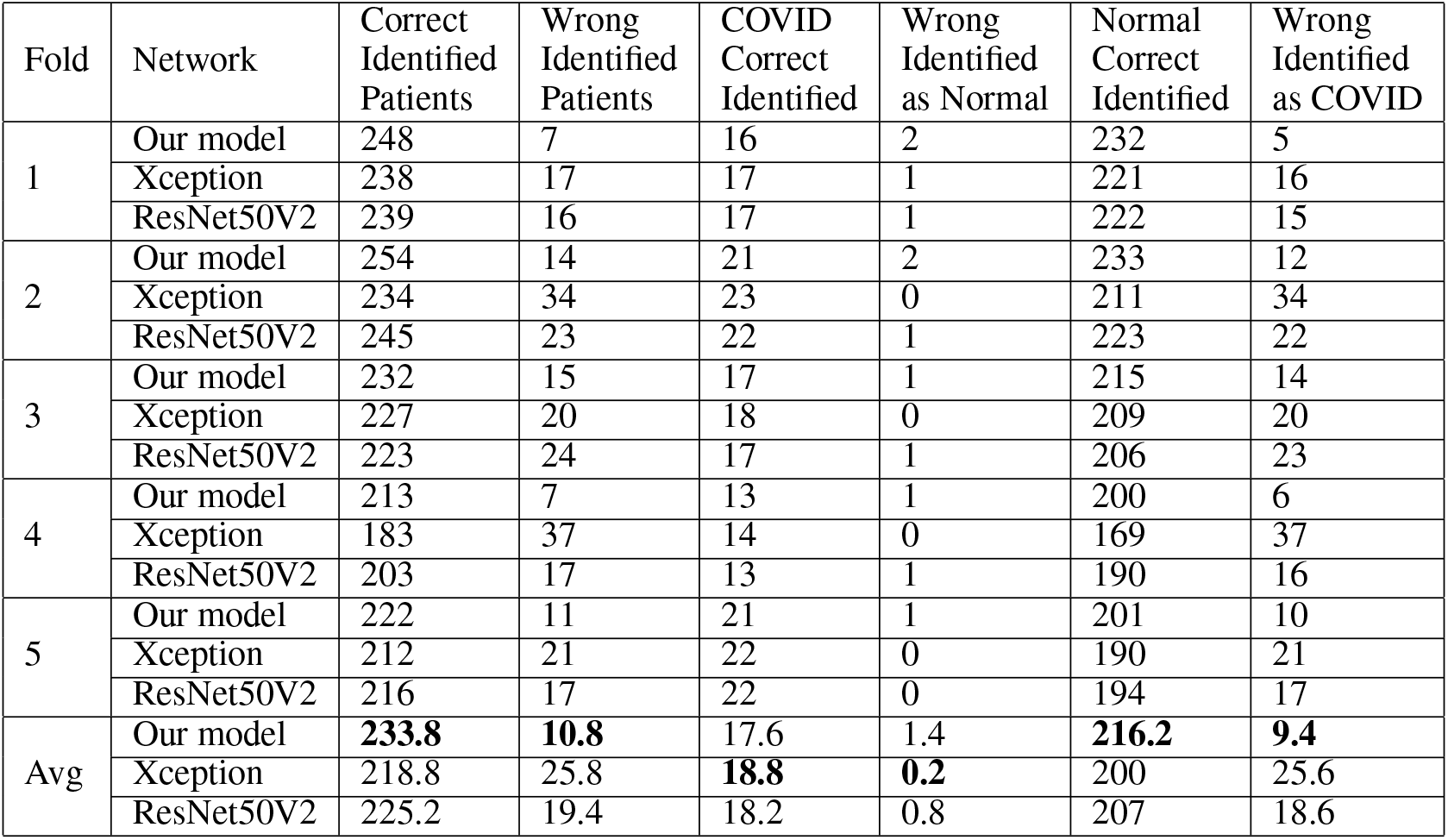
Patients identification results with the second threshold (0.1)

**Table 7:**
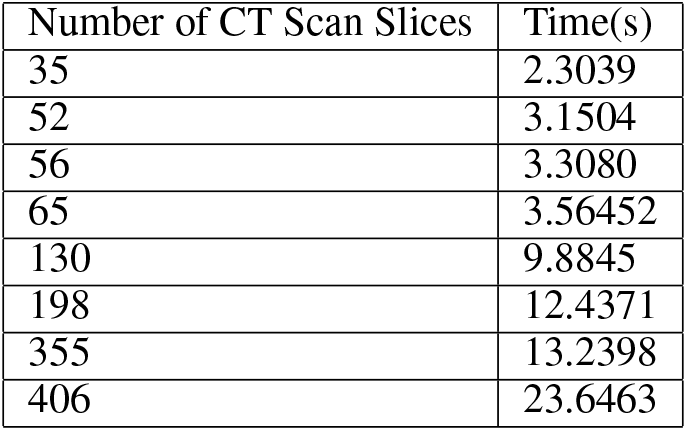
This table shows the fully automated system’s speed based on our model, for CT scans of different patients with various CT scan slices.

### 3.3 Feature Visualization

In this section, we aim to use the Grad-CAM algorithm [25] to visualize the extracted features of the network to determine the areas of infections and investigate the network’s correct performance.

By looking at Fig. 12 and comparing the normal and COVID images; it is visible that the network is classifying the images based on the infected areas. In the COVID-19 images, the highlighted features are around the infection areas, and in the normal images, as the network does not see any infections, the highlighted features are at the center showing the no infections have been found. Therefore the results can be trusted for medical diagnosis. Using the Grad-CAM algorithm can help the medical expert distinguish the CT scan images better and find the infections.

**Figure 12:**
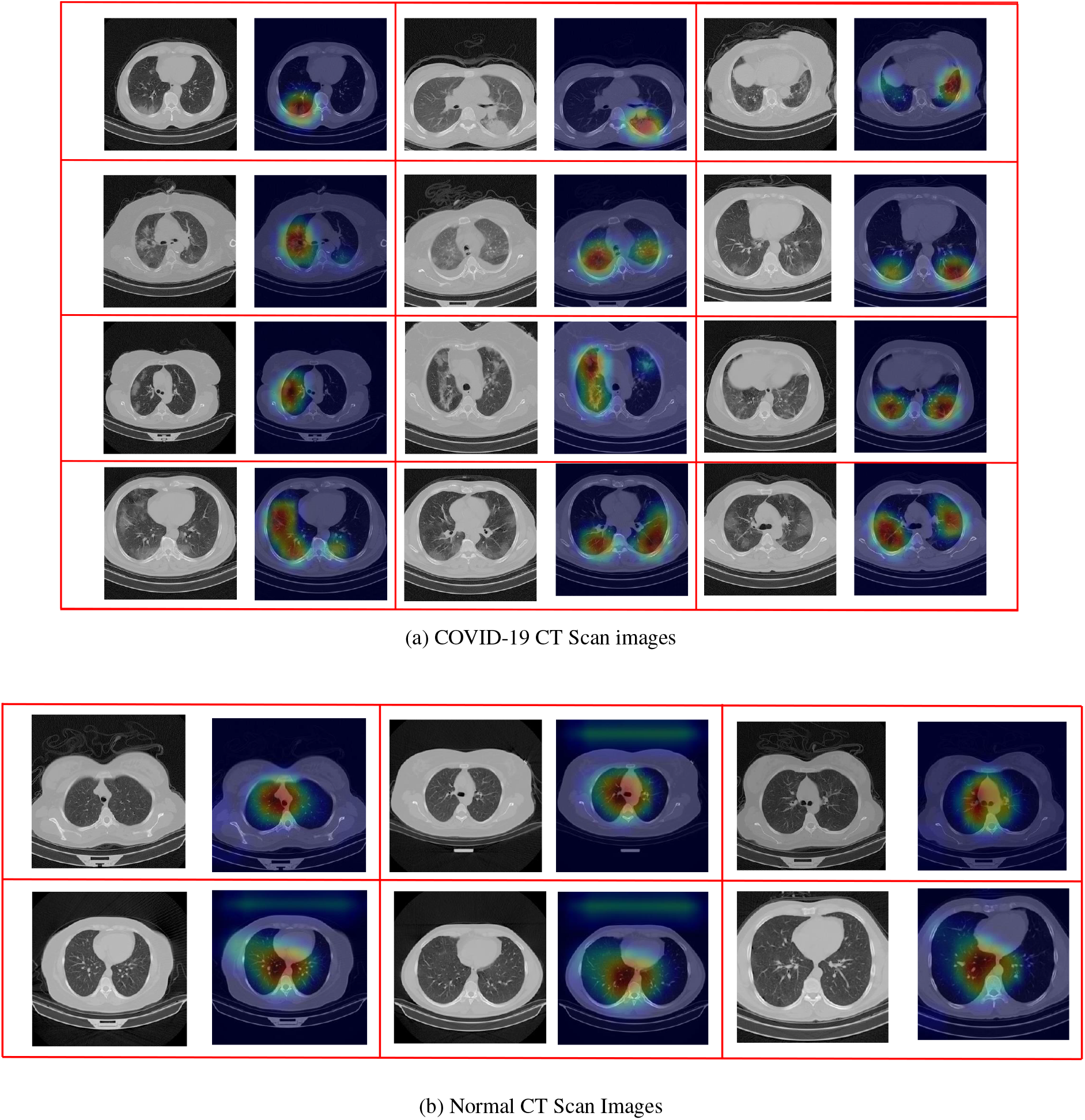
Visualized Features by the Grad-CAM algorithm to show that the network is operating correctly and indicate the infection regions in the COVID-19 CT Scan images

## 4 Discussion

Based on the results from table 8 and table 6, we understand that our proposed model made better accuracy. In the single image evaluating phase(table 8), the average results between five-folds show that our model achieved 98.49% overall accuracy and 94.96% sensitivity for the COVID-19 class. Xception evaluation results show 96.55 % overall accuracy and 98.02% COVID-19 sensitivity. Although Xception performed better in detecting COVID-19 patients, our model showed better results overall. This is because Xception sees any similar points as infections, and mistakenly identifies more normal images as COVID. But our model detects infection points carefully.

**Table 8:** Evaluation results for each network in each fold

| Fold | Network | Correct<br>Classified<br>Images | Wrong<br>Classified<br>Images | COVID<br>Correct<br>Classified | COVID<br>Not<br>Classified | Wrong<br>Classified<br>as COVID | Normal<br>Correct<br>Classified | Normal<br>Not<br>Classified | Wrong<br>Classified<br>as Normal | Overall<br>Accuracy | COVID<br>Accuracy | Normal<br>Accuracy | COVID<br>Sensitivity | Normal<br>Sensitivity | COVID<br>Specificity | Normal<br>Specificity | COVID<br>Precision | Normal<br>Precision |
| --- | --- | --- | --- | --- | --- | --- | --- | --- | --- | --- | --- | --- | --- | --- | --- | --- | --- | --- |
| 1 | Our model | 8214 | 108 | 436 | 26 | 82 | 7778 | 82 | 26 | 0.987 | 0.987 | 0.987 | 0.9437 | 0.9896 | 0.9896 | 0.9437 | 0.8417 | 0.9967 |
|  | Xception | 8165 | 157 | 456 | 6 | 151 | 7709 | 151 | 6 | 0.9811 | 0.9811 | 0.9811 | 0.987 | 0.9808 | 0.9808 | 0.987 | 0.7512 | 0.9992 |
|  | ResNet50V2 | 8169 | 153 | 453 | 9 | 144 | 7716 | 144 | 9 | 0.9816 | 0.9816 | 0.9816 | 0.9805 | 0.9817 | 0.9817 | 0.9805 | 0.7588 | 0.9988 |
| 2 | Our model | 8215 | 128 | 443 | 22 | 106 | 7772 | 106 | 22 | 0.9847 | 0.9847 | 0.9847 | 0.9527 | 0.9865 | 0.9865 | 0.9527 | 0.8069 | 0.9972 |
|  | Xception | 7921 | 422 | 458 | 7 | 415 | 7463 | 415 | 7 | 0.9494 | 0.9494 | 0.9494 | 0.9849 | 0.9473 | 0.9473 | 0.9849 | 0.5246 | 0.9991 |
|  | ResNet50V2 | 8109 | 234 | 456 | 9 | 225 | 7653 | 225 | 9 | 0.972 | 0.972 | 0.972 | 0.9806 | 0.9714 | 0.9714 | 0.9806 | 0.6696 | 0.9988 |
| 3 | Our model | 8143 | 186 | 427 | 19 | 167 | 7716 | 167 | 19 | 0.9777 | 0.9777 | 0.9777 | 0.9574 | 0.9788 | 0.9788 | 0.9574 | 0.7189 | 0.9975 |
|  | Xception | 8113 | 216 | 434 | 12 | 204 | 7679 | 204 | 12 | 0.9741 | 0.9741 | 0.9741 | 0.9731 | 0.9741 | 0.9741 | 0.9731 | 0.6803 | 0.9984 |
|  | ResNet50V2 | 8079 | 250 | 439 | 7 | 243 | 7640 | 243 | 7 | 0.97 | 0.97 | 0.97 | 0.9843 | 0.9692 | 0.9692 | 0.9843 | 0.6437 | 0.9991 |
| 4 | Our model | 8205 | 110 | 442 | 17 | 93 | 7763 | 93 | 17 | 0.9868 | 0.9868 | 0.9868 | 0.963 | 0.9882 | 0.9882 | 0.963 | 0.8262 | 0.9978 |
|  | Xception | 7854 | 461 | 449 | 10 | 451 | 7405 | 451 | 10 | 0.9446 | 0.9446 | 0.9446 | 0.9782 | 0.9426 | 0.9426 | 0.9782 | 0.4989 | 0.9987 |
|  | ResNet50V2 | 8166 | 149 | 439 | 20 | 129 | 7727 | 129 | 20 | 0.9821 | 0.9821 | 0.9821 | 0.9564 | 0.9836 | 0.9836 | 0.9564 | 0.7729 | 0.9974 |
| 5 | Our model | 8141 | 94 | 419 | 31 | 63 | 7722 | 63 | 31 | 0.9886 | 0.9886 | 0.9886 | 0.9311 | 0.9919 | 0.9919 | 0.9311 | 0.8693 | 0.996 |
|  | Xception | 8058 | 177 | 440 | 10 | 167 | 7618 | 167 | 10 | 0.9785 | 0.9785 | 0.9785 | 0.9778 | 0.9785 | 0.9785 | 0.9778 | 0.7249 | 0.9987 |
|  | ResNet50V2 | 7991 | 244 | 449 | 1 | 243 | 7542 | 243 | 1 | 0.9704 | 0.9704 | 0.9704 | 0.9978 | 0.9688 | 0.9688 | 0.9978 | 0.6488 | 0.9999 |
| Avg | Our model | 8183.6 | 125.2 | 433.4 | 23 | 102.2 | 7750.2 | 102.2 | 23 | **0.9849** | **0.9849** | **0.9849** | 0.9496 | **0.987** | **0.987** | 0.9496 | **0.8126** | 0.997 |
|  | Xception | 8022.2 | 286.6 | 447.4 | 9 | 277.6 | 7574.8 | 277.6 | 9 | 0.9655 | 0.9655 | 0.9655 | **0.9802** | 0.9647 | 0.9647 | **0.9802** | 0.636 | **0.9988** |
|  | ResNet50V2 | 8102.8 | 206 | 447.2 | 9.2 | 196.8 | 7655.6 | 196.8 | 9.2 | 0.9752 | 0.9752 | 0.9752 | 0.9799 | 0.9749 | 0.9749 | 0.9799 | 0.6988 | **0.9988** |

The reason COVID class precision is not very high, like accuracy or sensitivity, is because having an unbalanced test dataset. We had around 450 COVID-19 images and 7800 normal images for testing the network performance. Our model averagely classified 102 images from 7852 normal images as CODIV-19 wrongly, which is a good value, but because the number COVID-19 images in the test-set are much lower than normal test images, it made the COVID-19 precision around 81 percent. So this value of precision does not mean the network is performing poorly.

At the fully automated patient identification phase, we evaluated our model on almost 245 patients and 41892 images with different thicknesses. The average results between five folds in table 6 show that our model achieved the best results and approximately correctly classifies 234 persons from 245 persons, which is an acceptable value.

By referring to table 5, it can seen that our model performs patient identification more precisely than other networks. This shows the high ability of our developed model.

Fig. 12 also presents some of the classified images processed by the Grad-CAM algorithm to visualize the sensitive extracted features. Based on this figure, the system classifies the images by looking to correct points, and the results are trustworthy.

From table 7, it can be understood that the processing speed is good. As one CT scan image sequence has less than 100 images in most cases (when imaging thickness is between 2 to 6), this system can process them near 4 to 6 seconds. It can be seen from table 7 that some of the CT scan slices with a close difference differ in speed more than what is expected. The reason is that the CT scan selection algorithm may select different proper CT scan images from each of the CT image sequences. So the processing speed changes more than expectation.

What makes this work superior to other works is that this research can be implemented for real diagnosis because it is designed to work with the whole original CT scan data of patients with high accuracy and speed. Many of the previous works may claim to be close to the automated operation. Still, most of them have never evaluated their methods in real circumstances for automatic detection, such as detecting the condition of each patient on the whole original CT scans, evaluating their model on a large number of images, and not reporting network speed. Because of these matters, they can not be trusted in real COVID-19 diagnosis.

We hope that our shared dataset and codes can help other researchers improve AI models and use them for advanced medical diagnosis.

## 5 Conclusion

In this paper, we have proposed a fully automated system for COVID-19 detection from lung HRCT scans. We also introduced a new dataset containing 15589 images of normal persons and 48260 images belonging to patients with COVID-19. At first, we proposed an image processing algorithm to filter the proper images of the patients’ CT scans, which show inside the lung perfectly. This algorithm helps increase network accuracy and speed. At the next stage, we introduced a novel deep convolutional neural network for improving classification. This network can be used in many classification problems to improve accuracy.

We trained three different deep convolution networks for classifying the CT scan images into COVID-19 or normal. Our model, which utilizes ResNet50V2, a modified feature pyramid network, and the designed classification layers, achieved the best results. After training, we used the trained networks for running the fully automated COVID-19 identifier system.

We evaluated our system in two different ways: one on more than 7796 images and the other on almost 245 patients and 41892 images with different thicknesses. For single image classification (first evaluation way), our model showed 98.49% overall accuracy. Our model obtained the best results at the patient identification stage (second evaluation way) and correctly identified approximately 234 patients from 245 patients.

We also used the Grad-CAM algorithm to highlight the CT scan images’ infection areas and investigate the classification correctness. Based on the obtained results, it can be understood the proposed methods can improve COVID-19 detection and run fast enough for implementation in medical centers.

## Data Availability

Data and Codes are available at:
https://github.com/mr7495/COVID-CT-Code
https://github.com/mr7495/COVID-CTset

https://github.com/mr7495/COVID-CTset

## 6 Data availability

We have made our data available for public use in this address: (https://github.com/mr7495/COVID-CTset). The dataset is available in two parts: one is the raw data presented in three folders for each patient. The next part is the training, validation, and testing data in each fold. We hope that this dataset will be utilized for improving COVID-19 monitoring and detection in the coming researches.

## 7 Code availability

All the used codes for data analysis, training, validation, testing, and the trained networks are shared at (https://github.com/mr7495/COVID-CT-Code).

## Acknowledgment

We wish like to thank Negin radiology experts that helped us in proving the dataset. We also like to appreciate Google for providing free and powerful GPU on Colab servers and free space on Google Drive.

## Ethics Statement

We declare that this paper is original and has been read and approved by all named authors and that there are no other persons who satisfied the criteria for authorship but are not listed. We further confirm that all have approved the order of authors listed in the paper of us. All the patients’ shared data have been approved by Negin Radiology Medical Center located at Sari, Iran, under the supervision of its director(Dr.Sakhaei, radiology specialist) and Dr.Mahdi Hassanzadeh. It must be mentioned that to protect patients’ privacy; all the DICOM files have been converted to TIFF format files to remove the patients’ information.

## Footnotes

1 This dataset is shared at: https://github.com/mr7495/COVID-CTset

2 The details of each patient in our dataset are available at: https://github.com/mr7495/COVID-CTset/blob/master/Patient_details.csv

3 The thicknesses and other details of each patient in our dataset are available at: https://github.com/mr7495/COVID-CTset/blob/master/Patient_details.csv

